# Determinants of the incidence and mortality rates of COVID-19 during the first six months of the pandemic; a cross-country study

**DOI:** 10.1101/2021.01.21.21250226

**Authors:** Noha Asem, Ahmed Ramadan, Mohamed Hassany, Ramy Mohamed Ghazy, Mohamed Abdallah, Eman M. Gamal, Shaimaa Hassan, Nehal Kamal, Hala Zaid

## Abstract

COVID-19 pandemic raises an extraordinary challenge to the healthcare systems globally. The governments are taking key measures to constrain the corresponding health, social, and economic impacts, however, these measures vary depending on the nature of the crisis and country-specific circumstances.

**Objectives:** Considering different incidence and mortality rates across different countries, we aimed at explaining variance of these variables by performing accurate and precise multivariate analysis with aid of suitable predictors, accordingly, the model would proactively guide the governmental responses to the crisis.

**Methods:** Using linear and exponential time series analysis, this research aimed at studying the incidence and mortality rates of COVID-19 in 18 countries during the first six months of the pandemic, and further utilize multivariate techniques to explain the variance in monthly exponential growth rates of cases and deaths with aid of a set of different predictors: the recorded Google mobility trends towards six categories of places, daily average temperature, daily humidity, and key socioeconomic attributes of each country.

**Results:** The analysis showed that changes in mobility trends were the most significant predictors of the incidence and mortality rates, temperature and humidity were also significant but to a much lesser extent, on the other hand, the socioeconomic attributes did not contribute significantly to explaining different incidence and mortality rates across countries.

**Conclusion:** Changes in mobility trends across countries dramatically affected the incidence and mortality rates across different countries, thus, it might be used as a proxy measure of contact frequency.

## Introduction

The World Health Organization (WHO) estimates that approximately one-third (e.g., 20 million) of the annual deaths worldwide were attributed to infectious diseases. Furthermore, three of the 10-topped causes of deaths are lower respiratory tract infection, tuberculosis, and diarrheal disease, and many of these diseases can be prevented or treated by for as little as one dollar for head.^1^ The morbidity from infectious disease has increased during the past few decades and represents at least 70% of emerging infectious diseases (EID), which are a significant burden on global economic and public health. ^2, 3^ Emerging infectious diseases (EIDs) cause a substantial economic and public health burden in the world. ^4, 5^ The most likely causes of the emergence of EIDs are socioeconomic, environmental, and ecological factors. ^3-6^ It is postulated that the origins of EIDs are significantly correlated with socioeconomic, environmental and ecological factors that provide a clue for identifying regions where new EIDs are most likely to originate. ^6^ These factors also present a basis of risk for wildlife zoonotic and vector-borne EIDs originating at lower latitudes, where reporting effort is low ^6, 7^.

One of these EID is the novel corona virus (COVID-19) that was firstly identified in December 2019 in Wuhan city, in China. It resulted in unusual pneumonia after visiting animal market that sells poultry, fish and other animals to the public.^8^ This outbreak was soon reported to the World Health Organization (WHO). Due to this pandemic, the entire life has been changed. Millions of people have been infected (21,092,417), and hundreds of thousands have been deceased (757,738) by the novel corona virus.^9^Countries adopted different strategies to combat the spread of this pandemic. Many countries implemented the national lockdown policy that extended for different duration during daytime. Another strategy was social distancing, and panning going out of homes without waring facemasks. International travel from and to infected countries was another effective strategy used to face the spread of this pandemic. ^10, 11^

It is worthy to note that COVID-19 does not affect everyone in the same way, furthermore, it is not easy to understand the consequences and to predict how this pandemic affects differently various countries. There are several reasons that is why different population are affected by this pandemic in different ways. These conditions can include socioeconomic factors (income, population density, distribution of human population, urban and rural settings, education level, and lifestyle, the size of household, and homeowners & tenants), behavior factors (direct contact with domestic and wild animals, migration, social interactions), and environment factors (humidity, temperature, wind spread, climate change, deforestation, agricultural growth) ^12, 13^

This ecological study aimed at studying the effect of different sociodemographic criteria, environmental factors, and human behaviors that affect the transmission of COVID-19 and predict its virulence. Transmission rate was measured by daily incidence of new cases as reported by national health authority of studied countries, of new cases while virulence was assessed by the number of confirmed deaths.

## Methods

### Data collection

Eighteen countries were designated for analysis, representing different climates, socioeconomic status, population densities, and different counts of confirmed COVID-19 cases. Through the period from 1^st^ of January to 30^th^ of June 2020, the daily counts of COVID-19 confirmed cases and deaths for 18 countries were collected from https://ourworldindata.org/coronavirus-source-data, the daily average temperature and relative humidity of the capital cities of the selected countries were collected from https://www.wunderground.com/. Socioeconomic attributes of the selected countries were collected from https://ourworldindata.org/coronavirus-data, these attributes included the population density per Km^2^, Human Development Index (HDI), Per Capita Income (PCI), Gross Domestic Product (GDP) in (USD), internet coverage, density, surface (Km^2^), Population count, and proportion of the population aged 65 years or above, and proportion of the population suffering from diabetes mellitus. The recorded policy responses of the selected countries to COVID-19 were retrieved from the International Monetary Fund (https://www.imf.org/en/Topics/imf-and-covid19/Policy-Responses-to-COVID-19) accessed on 15^th^ of August 2020. For each of the selected countries (except for China and Iran), Google daily documentation on mobility trends across different categories of places such as retail and recreation, groceries and pharmacies, parks, transit stations, workplaces, and residential was retrieved. Google documentations was presented by percent change in mobility across the mentioned places compared to the baseline, which is the median value of mobility trends, for the corresponding day of the week, during the 5-week period Jan 3–Feb 6, 2020 ^(14)^. Google data was retrieved from https://www.google.com/covid19/mobility/index.html?hl=en.

### Statistical Analysis

Data analysis was conducted using SPSS software (version 25, IBM, USA). The Daily counts of confirmed cases and deaths for each country were split by month and were further linearly and exponentially modelled with time. The linear equation was based on the equation (y_1_ = a +βx_1_), where y is the confirmed cases or deaths for a given day of this month, the constant (a) is the number of cases at day 0 (the count before start of that month), β is the linear slope coefficient which is presenting the average daily increase in the count of confirmed cases or deaths within this month, and x is the day number of that month. On the other hand, the exponential equation is based on the equation (y1= a e^βx1^), where e^β^ is the average daily multiplying factor for increase or in other word, the exponential growth rate in a given month. The daily average temperatures of the capital cities were aggregated to monthly temperature for each country. Google documentation were handled using big data analytics with R software (version 3.5.3, http://cran.r-project.org; R Foundation for Statistical Computing, Vienna, Austria) to aggregate the data into the monthly mean percent change from the baseline across six categories of places for each of the selected countries.

The linear and exponential slope coefficients herein after referred to as linear growth rate and exponential growth rate, respectively. Non-parametric correlation between variables was assessed using Spearman rho’s test. The monthly exponential growth rates of confirmed cases and deaths were regressed versus the collected predictors for each country using mixed effects model with unstructured covariance. The highly correlated Variables was subjected to principal component analysis (PCA) with varimax rotation, and the extracted PCs were utilized for the regression models.

## Results

### Mathematical pattern of incidence and mortality at the start of the pandemic

Time series analysis was conducted for the monthly records of confirmed cases and deaths, both linear and exponential growth models were studied. The R^2^, constant, and β (Slope coefficient) of the linear and exponential models are presented in (**Supplementary material 1**). In January 2020, China was the only country that reported confirmed deaths with COVID-19, China, Australia, Canada, France, Germany, and the United States of America have reported confirmed cases with COVID-19 that are valid for statistical modeling. During January, the R^2^ of linear models for confirmed cases by time in most of the selected countries were higher than that of the corresponding R^2^ of the exponential model for the same data, in other words, the growing data for confirmed cases were better described with linear equations.

### Further Mathematical pattern of incidence and mortality

Starting from February 2020, and except for Egypt, Indonesia, and Brazil, all the selected countries started to report an increasing number of confirmed COVID-19 cases. Until June 2020, the R^2^ of the exponential models were occasionally relatively higher than that of the linear models for confirmed cases and deaths, while in most models they were nearly equal. The monthly exponential growth rates of confirmed cases and deaths were perfectly correlated (Spearman correlation coefficient = 0.945, *P* <0.01). For the 18 countries, the median monthly growth rates for confirmed cases and deaths are presented in **(Figure 1 and 2)**, respectively. We remarked that the increase in exponential growth in cases and deaths in a given month implies for an increase in linear growth in the next month, so the highest exponential growth in March implied for the highest linear growth in April 2020. This is because the exponential growth multiplies the counts over time, which in turn increases the absolute counts in the next month.

**Figure 1:**
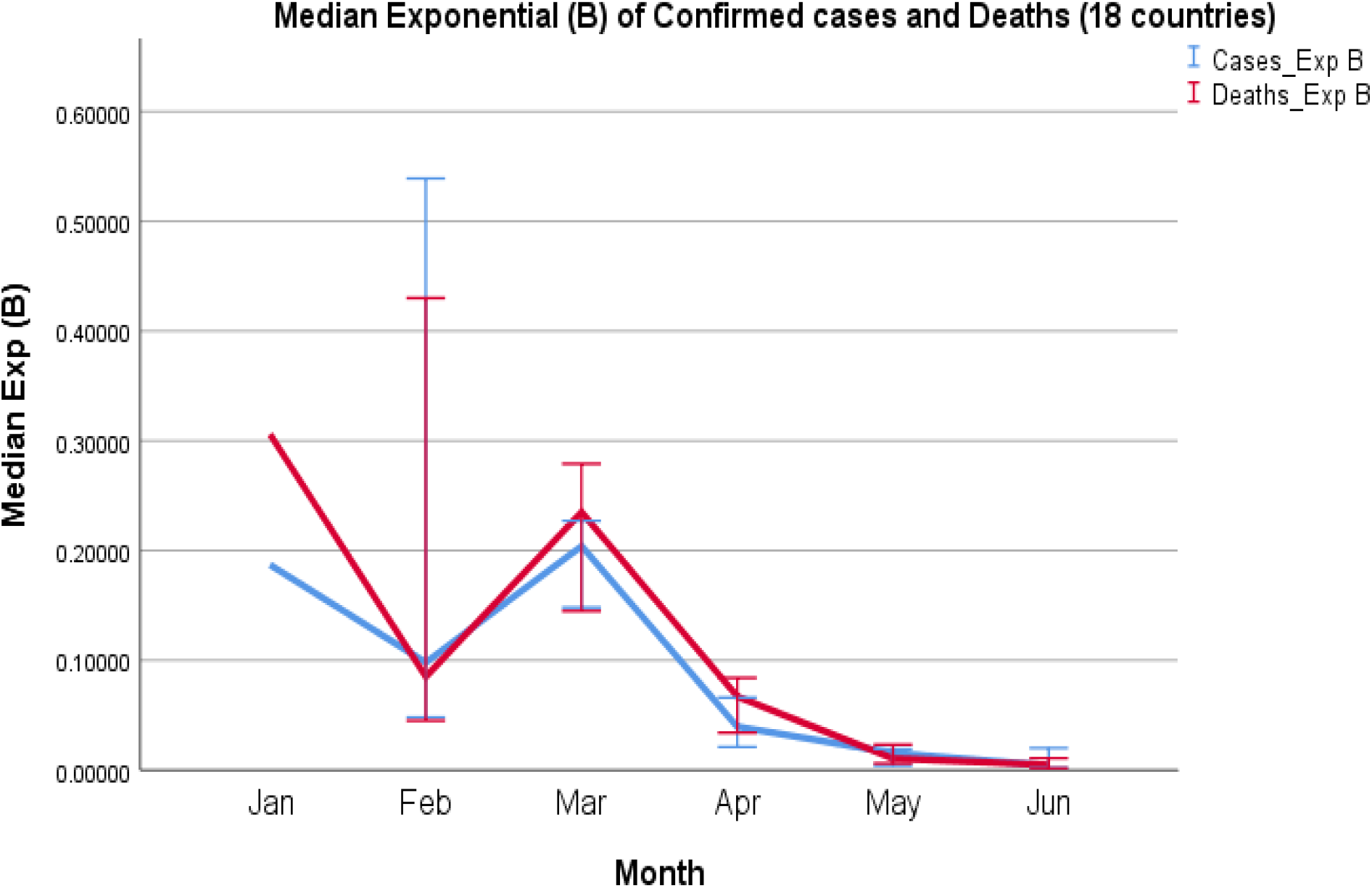
Median Monthly exponential growth rate of confirmed cases and deaths.

**Figure 2:**
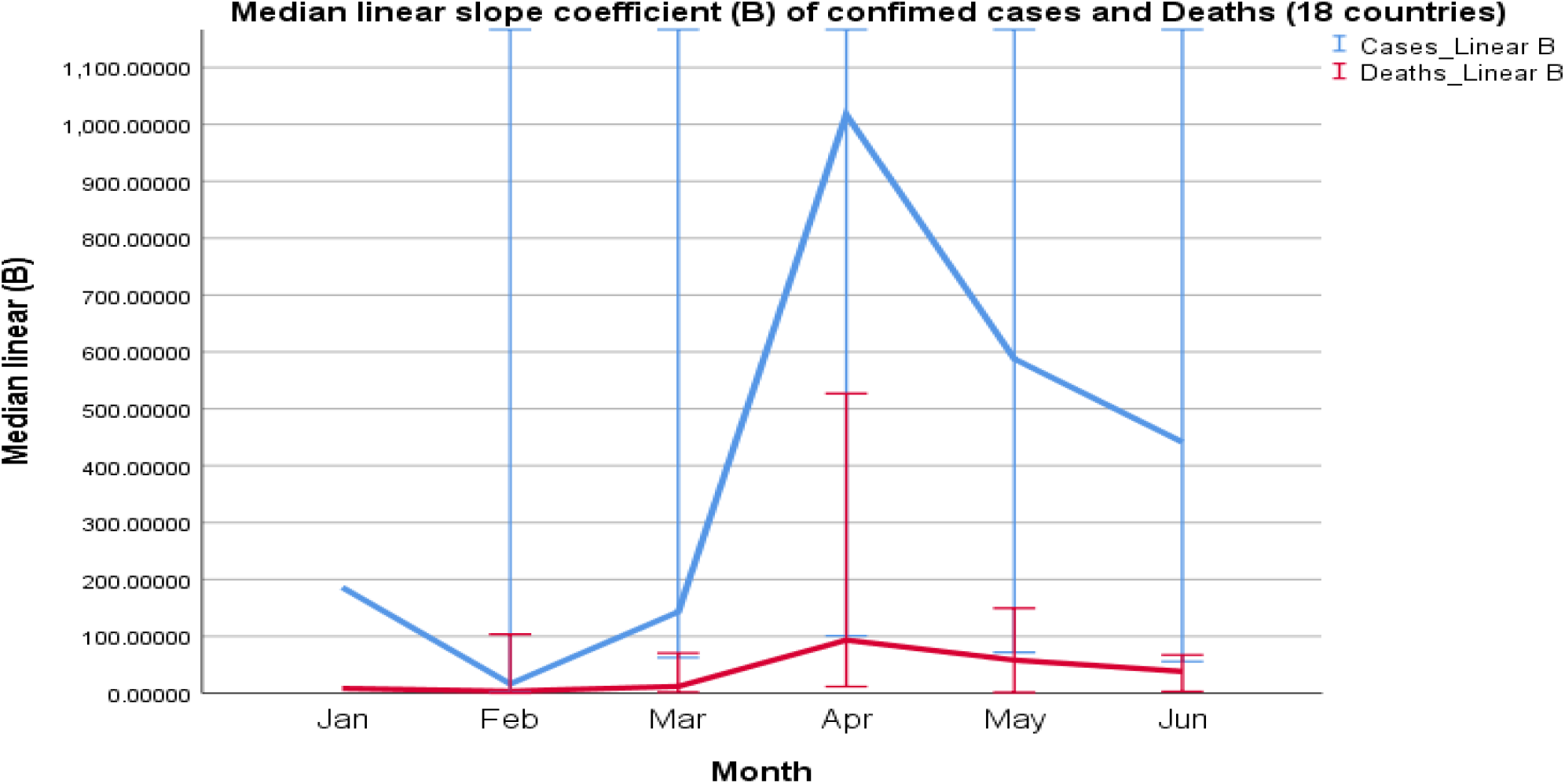
Median Monthly linear growth rate for confirmed cases and deaths.

### Pattern of mobility and its effect on incidence and mortality

The monthly change in mobility trends from baseline across different place categories are presented in **(Figure 3)**. In February 2020, mobility trends almost did not change from the baseline, then the median value started to decrease in March and April for all place categories except for mobility trends towards residential places. Afterwards, an inflection has occurred and the median values for mobility trends increased towards all place categories until the final timepoint in June 2020, in which most of median values were returned to the baseline. Interestingly, the median value of mobility trends toward parks, beaches and public gardens has even exceeded the baseline in June 2020 (+25%).

**Figure 3:**
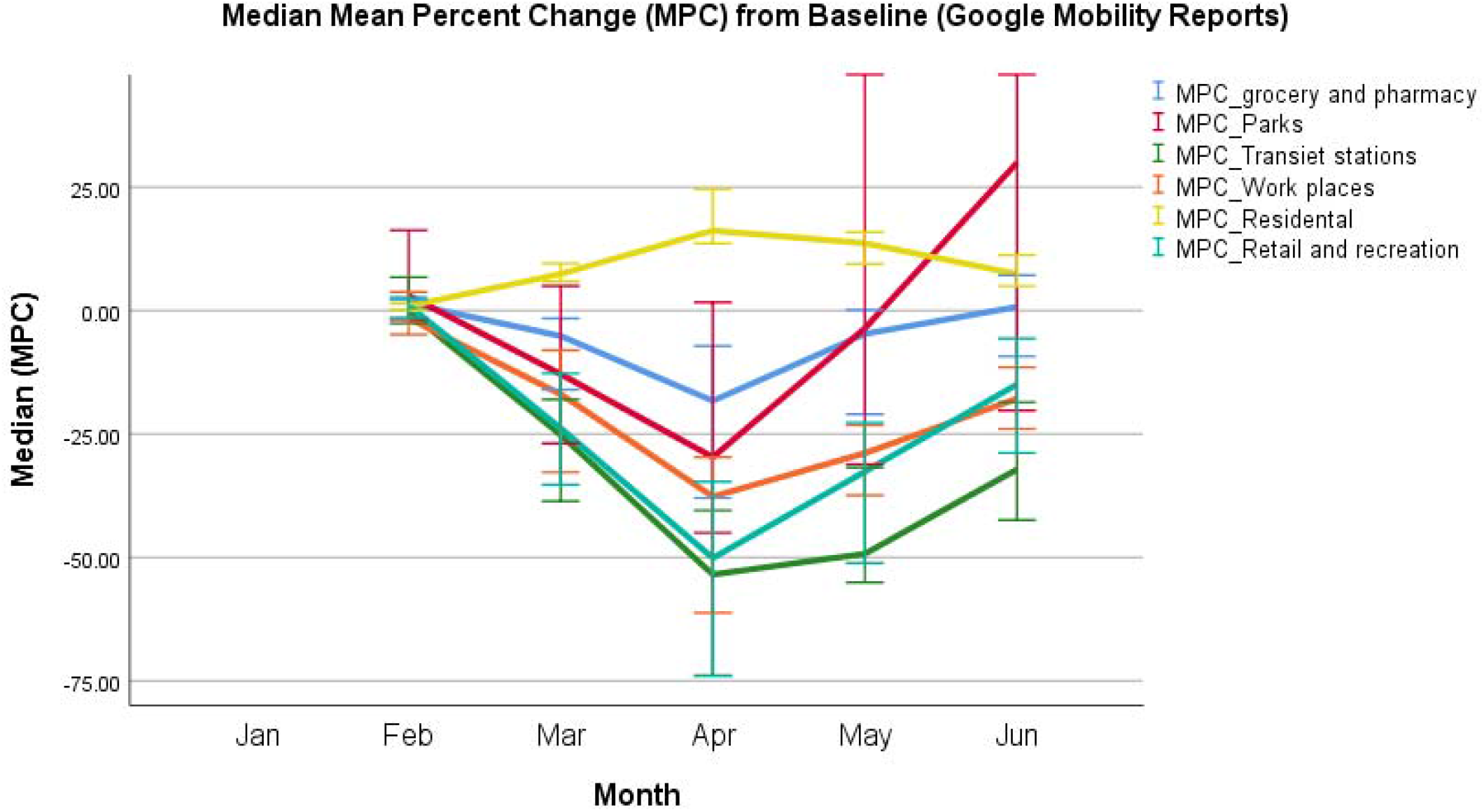
Median Change in mobility trends from baseline based on Google mobility reports.

It is worth noting that changes in mobility trends affected the exponential growth of incidence and deaths in the next month. So, we can notice a decrease in mobility trends from baseline in March, whose median exponential growth is the highest for cases and deaths, but we can further notice a decrease in exponential growth in the next month of April. This is because the confirmed cases usually get infected in a given month and been reported in the next month after a lag period of incubation and confirmation of infection. This remark was tested by assessing correlation between the exponential growth rates and mobility trends in the same month. The results are presented in **(Table 1)**, most of the correlations were week and insignificant, while when mobility trends were tested for correlation with the exponential growth rates of the next month, we got significant strong positive correlations for new cases and deaths **(Table 2)**.

**Table 1:** Correlations between changes in mobility trends toward six categories of places with exponential growth rates of cases and deaths of the same month.

| Correlations |  |  |  |  |  |  |  |  |  |  |
| --- | --- | --- | --- | --- | --- | --- | --- | --- | --- | --- |
|  |  |  | Cases_Exp<br>growth | Deaths_Exp<br>growth | grocery<br>and<br>pharmacy | Parks | Transient<br>stations | Workplaces | Residential | Retail<br>and<br>recreation |
| Spearman's<br>rho | Cases_Exp<br>growth | Correlation<br>Coefficient | 1.000 | .945** | -0.047 | -.319** | .244* | .311** | -0.187 | 0.078 |
|  |  | Sig. (2-tailed) | - | 0.000 | 0.684 | 0.005 | 0.032 | 0.006 | 0.103 | 0.502 |
|  | Deaths_Exp<br>growth | Correlation<br>Coefficient | .945** | 1.000 | -0.183 | -.345** | 0.077 | 0.070 | 0.048 | -0.121 |
|  |  | Sig. (2-tailed) | 0.000 | - | 0.142 | 0.005 | 0.541 | 0.576 | 0.699 | 0.335 |
| **. Correlation is significant at the 0.01 level (2-tailed). |  |  |  |  |  |  |  |  |  |  |
| *. Correlation is significant at the 0.05 level (2-tailed). |  |  |  |  |  |  |  |  |  |  |

**Table 2:**
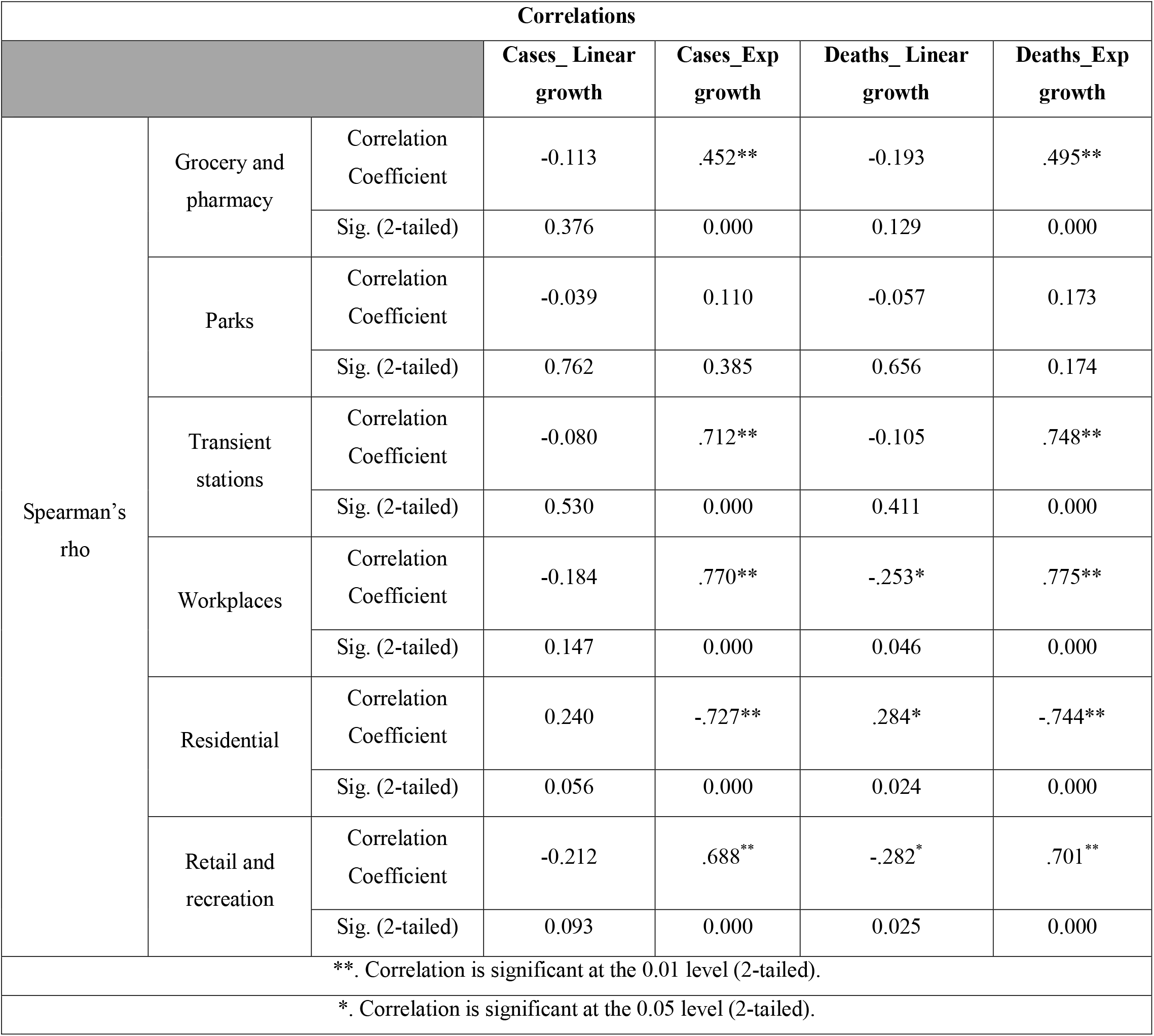
Correlations between changes in mobility trends with exponential growth rates of cases and deaths of the next month.

| Correlations |  |  |  |  |  |  |
| --- | --- | --- | --- | --- | --- | --- |
|  |  |  | Cases_ Linear growth | Cases_Exp growth | Deaths_ Linear growth | Deaths_Exp growth |
| Spearman's rho | Grocery and pharmacy | Correlation Coefficient | -0.113 | .452** | -0.193 | .495** |
|  |  | Sig. (2-tailed) | 0.376 | 0.000 | 0.129 | 0.000 |
|  | Parks | Correlation Coefficient | -0.039 | 0.110 | -0.057 | 0.173 |
|  |  | Sig. (2-tailed) | 0.762 | 0.385 | 0.656 | 0.174 |
|  | Transient stations | Correlation Coefficient | -0.080 | .712** | -0.105 | .748** |
|  |  | Sig. (2-tailed) | 0.530 | 0.000 | 0.411 | 0.000 |
|  | Workplaces | Correlation Coefficient | -0.184 | .770** | -.253* | .775** |
|  |  | Sig. (2-tailed) | 0.147 | 0.000 | 0.046 | 0.000 |
|  | Residential | Correlation Coefficient | 0.240 | -.727** | .284* | -.744** |
|  |  | Sig. (2-tailed) | 0.056 | 0.000 | 0.024 | 0.000 |
|  | Retail and recreation | Correlation Coefficient | -0.212 | .688** | -.282* | .701** |
|  |  | Sig. (2-tailed) | 0.093 | 0.000 | 0.025 | 0.000 |
| **. Correlation is significant at the 0.01 level (2-tailed). |  |  |  |  |  |  |
| *. Correlation is significant at the 0.05 level (2-tailed). |  |  |  |  |  |  |

### Effect of mobility on incidence of COVID-19

Monthly lagged exponential growth rates of cases showed intermediate positive correlation with mobility trends to glossaries and pharmacies (r = 0.452, P <0.01), and strong positive correlation with MPC in mobility trends to transit stations (r = 0.712, P<0.001), mobility trends to workplaces (r =0.77, P< 0.01), and mobility trends to retail and recreation (r=0.688, P<0.01), while it showed strong negative correlation with mobility trends to residential places (r= -0.727, P<0.001).

### Effect of mobility on mortality rate of COVID-19

Monthly lagged Exponential growth rates of deaths showed intermediate significant positive correlation with changes in mobility trends to groceries and pharmacies (r= 0.495, P<0.01), while it had a strong positive correlation with changes in mobility trends to retail and recreation places (r =0.7, P<0.01), changes in mobility trends to Work places (r=0.775, P<0.01), and changes in mobility trends to transit stations (r=0.748, P<0.01), and it showed a strong negative correlation with changes in mobility trends to Residential places (r= -0.744, P<0.01).

### Effect of temperature, humidity and sociodemographic factors on incidence cases and deaths

Monthly average temperatures of countries had a weak significant negative correlation with exponential growth rates of deaths in the next month (r = -0.25, P=0.038), while it did not significantly correlate with exponential growth rates of new cases (−0.08, P=0.49). Relative humidity was significantly correlated with the exponential growth rates of confirmed cases (r = 0.28, P=.017) and deaths (r=0.31, P =0.008) of the next month **(Table 3)**. On the other hand, we did not get any significant correlations upon testing the overall exponential growth rates for cases and deaths with the socioeconomic factors of the selected countries.

**Table 3:** Correlations between average temperature and humidity with exponential growth rates of cases and deaths of the next month.

| Correlations |  |  |  |  |  |  |
| --- | --- | --- | --- | --- | --- | --- |
|  |  |  | Cases_<br>Linear<br>growth | Cases_Exp<br>growth | Deaths_ Linear<br>growth | Deaths_Exp<br>growth |
| Spearman's<br>rho | Monthly<br>average<br>temp | Correlation<br>Coefficient | -0.131 | -0.083 | -0.172 | -.247* |
|  |  | Sig. (2-tailed) | 0.271 | 0.490 | 0.152 | 0.038 |
|  | Monthly<br>average<br>humidity | Correlation<br>Coefficient | -0.077 | .280* | -0.043 | .314** |
|  |  | Sig. (2-tailed) | 0.519 | 0.017 | 0.724 | 0.008 |
| **. Correlation is significant at the 0.01 level (2-tailed). |  |  |  |  |  |  |
| *. Correlation is significant at the 0.05 level (2-tailed). |  |  |  |  |  |  |

### Predictors of incidence and mortality

We modeled the exponential growth rates for cases and deaths with time (months), the principal component of mobility trends of the previous month, temperature of the previous month, humidity of the previous month, proportion of population above 65 years, diabetes prevalence, and PCI of each country as predictors.

### Model 1 (incidence rates)

The exponential growth rate of confirmed cases was significantly different across the tested four months (P<0.001). Moreover, the variable was significantly different in all pairwise comparisons of the tested timepoints (P<0.01). the intercept of the model was estimated at 0.16 (CI: 0.12-0.19, P<0.01), most of the decline in the dependent variable took place in April (−0.13, CI: -0.16 -0.10, P<0.01) and the decline continued until June (−0.16, CI: -0.19 -0.13, P<0.01) matching with the monthly decline in median values presented in **(Figure 1)**. mobility trends were a significant predictor with the highest estimate (0.00398, CI: 0.00159-0.00638, P=0.002) followed by temperature (0.000679, CI: 0.000169-0.001189, P=0.011), humidity (0.000249, CI: 0.000169-0.001189, P=0.000), and the proportion of patients above 65 years in each county (−0.000959, CI: -0.0016- -0.000026, P=0.012). The other predictors were insignificant. The model suggests that the exponential growth rate of COVID-19 confirmed cases would be fully controlled if mobility is decreased by about 40% of the baseline over 3 months (0.1552/0.00398) controlling for the other predictors. The model also suggests that one unit decline in mobility is equal in effect to about 6 degrees Celsius decline in temp (0.00398/0.000679) and about 16 degrees decline in relative humidity (0.00398/0.000249) **(Table 4)**.

**Table 4:** Estimates of fixed effects (Model 1)

| Parameter | Estimate | Std. Error | Df | t | Sig. | 95% Confidence Interval |  |
| --- | --- | --- | --- | --- | --- | --- | --- |
|  |  |  |  |  |  | Lower Bound | Upper Bound |
| Intercept | 0.155287 | 0.017119 | 26.127 | 9.071 | 0.000 | 0.120106 | 0.190468 |
| June | -0.163131 | 0.014233 | 16.828 | -11.461 | 0.000 | -0.193184 | -0.133078 |
| May | -0.153123 | 0.014151 | 17.062 | -10.821 | 0.000 | -0.182970 | -0.123275 |
| April | -0.129927 | 0.015765 | 16.189 | -8.242 | 0.000 | -0.163315 | -0.096539 |
| March | 0 | 0 |  |  |  |  |  |
| PC of mobility trends | 0.003987 | 0.001174 | 30.060 | 3.397 | 0.002 | 0.001590 | 0.006383 |
| Temp | 0.000679 | 0.000248 | 27.274 | 2.733 | 0.011 | 0.000169 | 0.001189 |
| Humidity | 0.000249 | 5.921562E-05 | 23.021 | 4.200 | 0.000 | 0.000126 | 0.000371 |
| proportion of population above 65 years | -0.000959 | 0.000318 | 11.136 | -3.019 | 0.012 | -0.001657 | -0.000261 |
| Diabetes prevalence | 0.000429 | 0.000514 | 14.015 | 0.835 | 0.418 | -0.000673 | 0.001532 |
| PCI | 2.479417E-08 | 1.632798E-07 | 14.691 | 0.152 | 0.881 | -3.238678E-07 | 3.734561E-07 |
| a. Dependent Variable: Cases_Exp growth. |  |  |  |  |  |  |  |
| b. Residual is weighted by Population Density (/km2). |  |  |  |  |  |  |  |

### Model 2 (mortality rates)

The exponential growth rate of deaths was significantly different across different months (P<0.01), and it was also significantly different in all pairwise comparisons of the tested timepoints (P<0.01). The intercept of the model was estimated at 0.20 (CI: 0.14-0.26, P<0.01), most of the decline in the dependent variable similarly took place in April (−0.14, CI: -0.19- -0.09, P<0.01) and the decline continued until June (−0.20, CI: -0.25 - 0.14, P<0.01) matching with the monthly decline in median values presented in **(Figure 1)**. Mobility trends had the highest estimate (0.0027, CI: 0.00001-0.0054, P=0.049) followed by temperature (0.0014, CI: 0.0009-0.0019, P<0.001), humidity (−0.0026, CI: 0.0041- -0.0001, P=0.000) and PCI of countries (−3×10^−7^, -5.72 x10^−7^- -9.57×10^−8^, P=0.009) **(Table 5)**.

**Table 5:** Estimates of fixed effects (Model 2)

| Estimates of Fixed Effects (Model 2) <sup>a, b</sup> |  |  |  |  |  |  |  |
| --- | --- | --- | --- | --- | --- | --- | --- |
| Parameter | Estimate | Std. Error | df | t | Sig. | 95% Confidence Interval |  |
|  |  |  |  |  |  | Lower Bound | Upper Bound |
| Intercept | 0.199858 | 0.027476 | 18.521 | 7.274 | 0.000 | 0.142249 | 0.257467 |
| June | -0.195938 | 0.024807 | 15.868 | -7.899 | 0.000 | -0.248561 | -0.143315 |
| May | -0.183183 | 0.024708 | 15.958 | -7.414 | 0.000 | -0.235573 | -0.130793 |
| April | -0.141371 | 0.022041 | 15.503 | -6.414 | 0.000 | -0.188218 | -0.094525 |
| March | 0 | 0 |  |  |  |  |  |
| PC of mobility trends | 0.002717 | 0.001309 | 23.276 | 2.076 | 0.049 | 1.127571E-05 | 0.005423 |
| Temp | 0.001426 | 0.000251 | 18.727 | 5.676 | 0.000 | 0.000900 | 0.001952 |
| Humidity | -0.000267 | 7.000827E-05 | 15.853 | -3.815 | 0.002 | -0.000416 | -0.000119 |
| proportion of population above 65 years | -8.677099E-05 | 0.000194 | 14.971 | -0.448 | 0.660 | -0.000499 | 0.000326 |
| Diabetes prevalence | 0.000404 | 0.000394 | 19.990 | 1.027 | 0.317 | -0.000417 | 0.001225 |
| PCI | -3.341040E-07 | 1.120112E-07 | 15.262 | -2.983 | 0.009 | -5.724931E-07 | -9.571477E-08 |
| a. Dependent Variable: Deaths_Exp growth. |  |  |  |  |  |  |  |
| b. Residual is weighted by Population Density (/km2). |  |  |  |  |  |  |  |

## Discussion

Unfortunately, the pandemic of COVID-19 is spreading very rapidly across the world causing huge number of deaths despite the relatively low case fatality ratio. Based on the world meter data on October 17, number of infected population approached 40 million while the number of deaths exceeded one million and one hundred thousand worldwide. ^14^ However, this is not the worst scenario, scientist warn the word from second wave of COVID-19 especially worldwide herd immunity is still away.^15^

In this research we tried to address different ecological factors that could affect the transmissibility and fatality of COVID-19. To the best of our knowledge, this study is the first to highlight the effect of different ecological factors like temperature, and humidity concurrently with studying other humanitarian factors like social mobility across different countries simultaneously. We randomly selected 18 countries with different disease statistics located at different sites on the world map. We remarked that the increase in Exp (β)of a given month implies for an increase in linear (β)in the next month, so the highest calculated median Exp (β)in March implied for the highest median linear (β)in April 2020. This is because the exponential growth multiplies the counts over time, which in turn increases the absolute counts in the next months. We further plotted the median monthly Exp (β)of confirmed cases and deaths of different continents.

### Exponential versus linear growth equation

In this research, we studied the both exponential growth and the linear pattern of COVID-19 spread within the earliest 6 months of COVID-19 pandemic. Similarly, in a study conducted by Comarova et al, ^16^ the pattern of growth was either exponential or power law growth. They further cleared that the pattern of spread of each country depended on the time when the pandemic took place; if the pandemic started early the pattern was exponential, while later pandemic following Italy was power law.

### Social mobility trends

Many countries adopt different strategies of national lockdown despite it damaging effect on economy and education. Mobility can be used as a proxy measure of contact frequency, so social mobility is drastically affecting the disease transmission. It is considered as one of the main control measures until an effective vaccine discovered. In the work presented the exponential growth of infection was significantly correlated with the social mobility pattern of the preceded month. This is explained by duration of incubation period of COVID-19 and reporting was recorded in the month followed. Moreover, Carteni et al,^17^ proclaimed that both the number of tested cases, proximity to the to the outbreak area and mobility of the citizen are the main predictors of COVID-transmission. One of the significant types of social mobility was trips; within 3 weeks trips which was significantly associated with transmission risk. Interestingly, this duration exceeded the containment duration which is 14 days. In this research, the effect of social mobility may be diluted due to inclusion of countries with different wealthy index. Weil et al, reported that wealthier communities had lower social mobility than poorer communities, furthermore, the direction of mobility was more toward the least crowded areas before the pandemic than the more crowded ones and vice versa in poorer communities. ^18^ In United Kingdom, reduction of social mobility to 57.3% to 65.9% of the pre-lockdown situation would decrease the reproduction rate below 1 which means pandemic control. Reports noted that social mobility contributes to 80% of disease transmissibility. ^19^ Similarly, Badr et al, ^20^ reported the Pearson correlation between COVID-19 transmission and social mobility exceeded 0.7 in 25 states in USA. In this research, the effect of social mobility may be diluted due to inclusion of countries with different wealthy index and studying other important environmental factors. Weil et al, reported that wealthier communities had lower social mobility than poorer communities, furthermore, the direction of mobility was more toward the least crowded areas before the pandemic than the more crowded ones and vice versa in poorer communities. ^18^

In the current study, social mobility was revealed to be the most important determinant of the incidence and virulence of COVID-19 infections. This was simultaneously controlled by the presence of the other different determinants in the same model. The mobility patterns across the studied countries were a reflection for the nationwide decisions taken for controlling the pandemic. Most of these decisions were effective by March 2020, responding to the dramatic increase in confirmed cases and deaths. Accordingly, social mobility was decreased which in turn decreased the incidence of the disease. However, due to several causalities, most countries decided to lessen the precautionary measures and made a gradual re-opening. Most of the re-opening decisions were effective by May and June 2020. Interestingly, we remarked an inflection in the social mobility graphs in April 2020, thus we are worried about a potential correlation with a second wave of the pandemic. Interestingly, the model suggests that 40% decrease in social mobility would control the incidence of the infections in 3 months, taking into considerations that the model was utilizing data from March to June 2020, during which the toughest decisions were taken by the governments.

### Temperature and Humidity

A questionable issue arose during this pandemic, whether climatic or demographic characteristics enable a more significant transmission of the virus. Environmental temperature was thought to affect the virus survival on surfaces consequently it affects viral transmissibility. The results clearly support the first reported statistically significant relationship of negative correlation between the average environmental temperature and exponential growth rates of the infected cases.^21^ Mecenas et al, ^22^ reviewed the results of 517 published research articles that evaluated the effect of humidity and temperature on the viral spread. Only 17 articles fulfilled the inclusion criteria, they concluded that hot and wet weather significantly affect the viral transmission. On the same line, Prata et al ^23^ demonstrated that there is more than 4.8% reduction in the cumulative incidence of newly diagnosed cases with each rise in temperature by 1 °C. Interestingly, Adly, ^24^ demonstrated that the optimal temperature for virus transmission was between 13-24 ° C based on analysis of data from March to November 2020. In this research, we also remarked that temperature and humidity had a significant determinant on the incidence of COVID-19, however, the effect was trivial compared to that of social mobility

### Socioeconomic factors

Since the beginning of the pandemic, there was no clue on the contribution of countries’ socioeconomic attributes to the propagation of COVID-19 cases. To the best of our knowledge, no previous studies investigated these factors as determinants of transmission. In the current study, and despite showing non-significant contribution to the incidence and virulence of the infection, these attributes remain of interest for the across-country studies for controlling the other investigated attributes.

### Limitation and strength

To the best of our knowledge, this research is the first to address the pattern of the spread either linear or exponential, it modelled the effect of social mobility environmental temperature, and humidity simultaneously, we did not include other important determinant of health like income, health care facilities, access to health systems, comorbidities and extra. An important point of limitation was that case definition was variable across different countries. Furthermore, case definition of suspected case or confirmed case may vary across time in the same country. Absolutely this may affect the number of reported cases and the same problem can be encountered for number of deaths. These facts may affect the external validity of our models.

## Conclusion

Data-driven suppositions can efficiently and proactively guide the governmental measures taken to lessen the social, health, and economic impacts of COVID-19 pandemic. During the first six months of the pandemic, the multivariate analysis showed that the changes in mobility trends across countries dramatically affected the incidence and mortality rates across different countries, thus, it might be used as a proxy measure of contact frequency. Studying a set of different predictors in our models makes the estimates more accurate and precise, because the contribution of each predictor is controlled for the contribution of the other predictors in the models.

## Data Availability

All data sources are explicitly stated. Datasets are available upon request

## Declarations

### Ethics approval and consent to participate

Not applicable

### Consent for publication

Not applicable

### Availability of data and materials

The datasets used and/or analyzed during the current study are available from the corresponding author on reasonable request.

## Acknowledgement

Not applicable

## References

1. Word Health Organization. World Health Report. https://www.who.int/whr/1996/media_centre/press_release/en/

2. Morens DM, Folkers GK, Fauci AS. The challenge of emerging and re-emerging infectious diseases. Nature. 2004;430(6996):242–249.

3. Dye C. After 2015: infectious diseases in a new era of health and development. Philosophical Transactions of the Royal Society B: Biological Sciences. 2014;369(1645):20130426.

4. Halliday JE, Hampson K, Hanley N, et al. Driving improvements in emerging disease surveillance through locally relevant capacity strengthening. Science. 2017;357(6347):146–148.

5. Binder S, Levitt AM, Sacks JJ, Hughes JM. Emerging infectious diseases: public health issues for the 21st century. Science. 1999;284(5418):1311–1313.

6. Woolhouse ME, Gowtage-Sequeria S. Host range and emerging and reemerging pathogens. Emerging infectious diseases. 2005;11(12):1842.

7. Jones KE, Patel NG, Levy MA, et al. Global trends in emerging infectious diseases. Nature. 2008;451(7181):990–993.

8. Xu Z, Shi L, Wang Y, et al. Pathological findings of COVID-19 associated with acute respiratory distress syndrome. Lancet Respir Med. 2020. DOI: https://doiorg/101016/S2213-2600(20). 2020;

9. Worldmeter. Corona Virus. Accessed 14 August, 2020. https://www.worldometers.info/coronavirus/

10. Cheng VC-C, Wong S-C, Chuang VW-M, et al. The role of community-wide wearing of face mask for control of coronavirus disease 2019 (COVID-19) epidemic due to SARS-CoV-2. J Infect. 2020;81(1):107–114. doi:10.1016/j.jinf.2020.04.024

11. Organization WH. COVID□19 STRATEGY UPDATE. 2020. file:///C:/Users/ramy/Downloads/covid-strategy-update-14april2020.pdf

12. Dehghani R, Kassiri H. A brief review on the possible role of houseflies and cockroaches in the mechanical transmission of coronavirus disease 2019 (COVID-19). Archives of Clinical Infectious Diseases. 2020;15(COVID-19)

13. Barratt R, Shaban RZ, Gilbert GL. Clinician perceptions of respiratory infection risk; a rationale for research into mask use in routine practice. Infection, disease & health. 2019;24(3):169–176.

14. Worldmeter. COVID-19 CORONAVIRUS PANDEMIC. Accessed October 17, 2020. https://www.worldometers.info/coronavirus/

15. Wise J. Covid-19: Risk of second wave is very real, say researchers. BMJ: British Medical Journal (Online). 2020;369

16. Komarova NL, Schang LM, Wodarz D. Patterns of the COVID-19 pandemic spread around the world: exponential versus power laws. Journal of the Royal Society Interface. 2020;17(170):20200518.

17. Cartenì A, Di Francesco L, Martino M. How mobility habits influenced the spread of the COVID-19 pandemic: Results from the Italian case study. Science of The Total Environment. 2020/11/01/ 2020;741:140489. doi:https://doi.org/10.1016/j.scitotenv.2020.140489

18. Weill JA, Stigler M, Deschenes O, Springborn MR. Social distancing responses to COVID-19 emergency declarations strongly differentiated by income. Proceedings of the National Academy of Sciences. 2020;117(33):19658. doi:10.1073/pnas.2009412117

19. Sussex Uo. Risk of return to exponential growth in Coronavirus cases if UK mobility not kept below half of pre-lockdown levels. Accessed september, 3, 2020. http://www.sussex.ac.uk/broadcast/read/52206

20. Badr HS, D. H, Marshall M, Dong E, Squire MM, Gardner LM. Association between mobility patterns and COVID-19 transmission in the USA: a mathematical modelling study. The Lancet Infectious Diseases. 2020;

21. Livadiotis G. Statistical analysis of the impact of environmental temperature on the exponential growth rate of cases infected by COVID-19. PLoS One. 2020;15(5):e0233875–e0233875. doi:10.1371/journal.pone.0233875

22. Mecenas P, Bastos R, Vallinoto A, Normando D. Effects of temperature and humidity on the spread of COVID-19: A systematic review. medRxiv. 2020;

23. Prata DN, Rodrigues W, Bermejo PH. Temperature significantly changes COVID-19 transmission in (sub)tropical cities of Brazil. Science of The Total Environment. 2020/08/10/ 2020;729:138862. doi:https://doi.org/10.1016/j.scitotenv.2020.138862

24. Anis A. The Effect of Temperature Upon Transmission of COVID-19: Australia And Egypt Case Study. Available at SSRN 3567639. 2020;

